# The prevalence, trends and heterogeneity in maternal smoking around birth between the 1930s and 1970s

**DOI:** 10.1101/2023.10.09.23295827

**Authors:** Stephanie von Hinke, Jonathan James, Emil Sorensen, Hans H. Sievertsen, Nicolai Vitt

## Abstract

This paper shows the prevalence, trends and heterogeneity in maternal smoking around birth in the United Kingdom, focusing on the war and post-war reconstruction period in which there exists surprisingly little systematic data on (maternal) smoking behaviours. Within this context, we highlight relevant events, the release of new information about the harms of smoking, and changes in (government) policy aimed at reducing smoking prevalence. We show stark changes in smoking prevalence over a 30-year period, highlight the onset of the social gradient in smoking, as well as genetic heterogeneities in smoking trends.

## 1 Introduction

The offspring health impacts of prenatal smoking have been a topic of research for over a century. Animal studies from the early to mid 1900s showed a reduction in fetal growth, birth weight, and red blood cell counts after prenatal exposure to tobacco smoke and nicotine, as well as an increase in prenatal and neonatal deaths (see e.g., Guillain and Gy, 1907; Fleig, 1908; Pechstein and Reynolds, 1937; Schoeneck, 1941; Willson, 1942). Others noted similar growth restrictions for tobacco exposure in early life (see e.g., Richon and Perrin, 1908; Fleig, 1908), suggesting this to be driven by nicotine inhibiting lactation (e.g., Hatcher and Crosby, 1927) or by the secretion of nicotine into milk (e.g., Willson, 1942).

The first research papers that showed the detrimental impacts of tobacco on *human* health were published in the late 1940s and early 1950s (Kennaway and Kennaway, 1947; Doll and Hill, 1950), soon followed by research focusing on the impacts of *prenatal* smoking on offspring outcomes around birth. These studies showed that the results from animal studies extended to humans, as maternal smoking was associated with fetal growth restrictions and an increase in the likelihood of infants being premature, with stronger associations for heavier smokers (Simpson, 1957). These associations were robust to controlling for a range of confounders such as maternal age, parity and social circumstances (Lowe, 1959). Since then, many subsequent studies have shown the adverse impacts of *in utero* exposure to tobacco and cigarette smoke on outcomes around birth (e.g., birth weight; Rantakallio, 1978b; Walker et al., 2009; Yang et al., 2020; Pereira et al., 2022) but also the impacts of prenatal smoking on outcomes in the longer run (e.g., hospital admissions, childhood height, educational outcomes, child behaviour; Butler and Goldstein, 1973; Rantakallio, 1978a; Dolan et al., 2016).^1^ While the harmful effects of smoking during pregnancy are well-known today and maternal smoking rates are lower than decades ago (see e.g. Fertig, 2010), we have limited evidence on how the prevalence of maternal smoking developed during the period when much of the evidence was established, as well as how maternal smoking rates varied across different groups in society.

This paper explores the prevalence, trends and heterogeneity in maternal smoking around birth in the United Kingdom. We focus on women who had children between the late 1930s and early 1970s. Between 2006 and 2010, these *children* were asked whether their mother smoked around the time of their birth, allowing us to get an interesting insight into maternal smoking during the war and post-war reconstruction period. We also highlight two types of heterogeneity in the prevalence and trends of smoking behaviour during this time: by social class and by individuals’ genetic variation.

Perhaps surprisingly, very little systematic individual-level data exists about the prevalence of smoking in the UK (especially *female* or *maternal* smoking) during this period, its heterogeneity, and its potential drivers. An exception is Forster and Jones (2001) who use retrospective individual-level data on smoking, or Jones (1989a), who model aggregate smoking data between 1954 and 1986). Instead, most systematic data collection on smoking focuses on the post-1970s period. This is perhaps unexpected, since there was much research on the potential impacts of smoking (e.g., lung cancer), and even some on the impact of maternal smoking on offspring (see above). These studies, however, tended to collect their own data using specific population subgroups (e.g., hospital patients) or small samples. This means that relatively little is known about the prevalence, trends and heterogeneity in maternal smoking during this period. For example, although Wald and Nicolaides-Bouman (1991) provide “a comprehensive description of smoking in the United Kingdom by amalgamating the published and unpublished data from various sources”, with one chapter focusing solely on smoking during pregnancy, it merely highlights the prevalence of prenatal smoking in a limited and selected number years from 1958 to 1986. It also highlights the social gradient in maternal smoking in the 1980s, but focuses on a few years and does not show any statistics for the pre 1958 period.

We use the UK Biobank, a prospective cohort study that focuses on the health and well-being of over half a million individuals living in the UK. Participants are born between the late 1930s and early 1970s, and the data record rich information on participants’ health and economic outcomes in older age. Individuals are also asked whether their mother smoked around their birth. This is the main information that we use in this paper. We also highlight relevant events, the release of new information about the harms of smoking, and changes in (government) policy around this time period that may explain the (change in) trends and gradients that we observe. Note, however, that because of the many changes in society and government policy at the time, some happening relatively close together, we do not attempt to identify the impact of these separate events. The chapter is therefore merely descriptive, showing the trends and gradients that we observe over this period.

Although this is a large UK study, it is important to note that it is not representative of the population in the United Kingdom. On average, UK Biobank participants are healthier and wealthier (Fry et al., 2017), implying that we cannot necessarily generalize our findings to the full UK population, and our descriptive statistics and trends capture variation in maternal smoking among a more select group of women in the United Kingdom. Nevertheless, it allows us to describe systematic trends in maternal prenatal smoking during the second half of the 20th century for a group of slightly wealthier and healthier individuals; something that has not been possible until now. Furthermore, merging in data on occupational socioeconomic status (SES) at the local area level from the 1951 Census allows us to explore social gradients in prenatal smoking among this group as well as how this changed over time. Finally, exploiting the genetic data available in the UK Biobank allows us to explore genetic heterogeneity in these trends. This sheds further light on the variation in maternal smoking and *when* as well as *how* this changes over time.

The prevalence, trends and heterogeneity in smoking during this time period are important and interesting for multiple reasons. In addition to it shedding light on societal norms and values during this period, it allows us to better understand societal changes in smoking perceptions, and how policy may have helped shape these. Indeed, understanding trends in maternal smoking rates – and heterogeneity thereof – within the societal context of the time may help us learn more about *how* individuals respond to information and (government) policy, as well as *who* is most likely to respond. Furthermore, because the early life environment can have life-long and irreversible impacts on offspring from birth to older age^2^, a better understanding of the trends in maternal smoking during our period of observation may explain some of the variation in outcomes in adulthood and older age that we observe nowadays.

Our paper speaks to the literature on the determinants of smoking, such as those identifying tax elasticities for starting and quitting smoking (see e.g., Forster and Jones, 2001, who investigate smoking in general (rather than *maternal* smoking) during this time period), studies that explore the determinants of starting and quitting (see e.g., Yen and Jones, 1996; Jones, 1994; Etiĺe and Jones, 2011), as well as the best ways of modelling smoking decisions (see e.g., Jones, 1989b,c, 1992; Jones, 1989a; Jones and Labeaga, 2003). Our paper, however, is descriptive in nature. Rather than identifying the determinants of smoking behaviour or highlighting methodological issues that are relevant in modelling smoking behaviour, we provide a thorough description of maternal smoking across a thirty-year period, highlighting relevant social, contextual and legislative changes for this generation of mothers.

The rest of this chapter is structured as follows: Section 2 provides a background to the chapter, discussing the history of smoking regulation and other (government) policy that may affect smoking take-up. The data is described in Section 3, showing the general trends in maternal smoking during this time period, with the heterogeneity analysis shown in Section 4. We conclude in Section 5.

## 2 Background

Tobacco was one of the few items that was never rationed during the Second World War, since the government thought doing so would be bad for morale and it would be in conflict with tobacco’s revenue-raising function (Zweiniger-Bargielowska, 2000). However, demand far exceeded supply, leading to a well-functioning black market, where demand was subdued due to the high price, with some anecdotal evidence suggesting that tobacco was generally sold on the black market for three times its normal price.

Tobacco taxes at this time were relatively low. In April 1947, however, the then-Chancellor of the Exchequer, Hugh Dalton, dramatically increased the tobacco customs duty. He stated that he would raise this “from tomorrow by about 50 per cent (…) The effect of this increase will be that the price of a packet of 20 cigarettes will be raised from 2s. 4d. to 3s. 4d.”^3^ The rationale for the increase was not to improve health or to increase tax revenue. Rather, the concern was that the vast majority of tobacco consumed was imported from the United States and “to satisfy this insatiable demand, we are drawing heavily and improvidently on the dollars which we earn with our exports. (…) I regard the saving of dollars as much more important than an increase in the revenue in this connection.” Indeed, the budget was dubbed a “save the dollars” budget.^4^ The tax rise implied an overnight 43 percent increase in the price of a pack of 20 cigarettes.^5^

In 1948, however, health was on the minds of the Labour administration. On 5th July the National Health Service (NHS) was established, following the 1942 Beveridge report which set out the degree to which there were social and health inequalities in the UK. The NHS had three main goals: free provision of healthcare, access based on clinical need, and equalisation of access to medical services. The NHS was funded through general taxation and was free at the point of use. Before the establishment of the NHS, healthcare was mainly provided through private doctors and hospitals, and limited access to free healthcare was available through voluntary hospitals and local authority-run hospitals under the Poor Law. Local authorities were in charge of a number of public health programmes including ante-natal clinics and domiciliary midwifery. Once the NHS was introduced, hospitals were taken into public ownership and general practitioners (GPs) became contractors that were paid a set fee per treatment. Local authorities remained in charge of family health services.

The NHS dramatically changed women’s ability to access healthcare. Lührmann and Wilson (2018) document the impact that the introduction of the NHS had on health outcomes. In particular, they find a drop in infant mortality for the affected cohorts. The increase in access to both pre and post-natal care would have been greater for those who were likely to be uninsured prior to the introduction of the NHS. Indeed, Lührmann and Wilson (2018) find larger mortality reductions in areas with lower expected levels of health insurance (proxied by the proportion of illegitimate births). While ante-natal clinics remained in the control of local authorities, women now had free access to general practitioners (GPs) and – as part of that – maternity services.^6^ This greater access to primary care may have potentially provided women with greater access to information about the dangers of smoking via their GP as that information diffused through the medical community.

It had been long thought that smoking was dangerous and deleterious to health. The first anti-smoking literature was published in 1604 by King James I of England. In his “Counterblaste to Tobacco” he described smoking as “..a custom loathsome to the eye, hateful to the nose, harmful to the brain, dangerous to the lungs” (*King James I* 1604). Then the RCP met in 1605 to discuss the King’s pamphlet, however, they dismissed his views (ASH, n.d.). It was not until some 300 years later that the medical and academic community began to catch up with the King.

The key publication showing the adverse effects of smoking was that by Doll and Hill (1950). Using a case-control study that examined smoking-habits of lung cancer patients, they showed that those who smoked were 20 times more likely to develop lung cancer compared to non-smokers. They also found that the risk of lung cancer increased with the number of cigarettes smoked per day and the duration of smoking. Simultaneously, in the US, Wynder and Graham (1950) were working on the same issue and discovered the same pattern. Ronald Fisher was very critical of this work. He was concerned that the estimates reflected correlations rather than causation and was strongly against antismoking publicity which he dubbed propaganda (Fisher, 1959). Over the next decade further work established the link between smoking and carcinoma of the lung as well as other diseases and made attempts to establish that this relationship was causal. Doll and Hill extended their work, starting ‘The British Doctors Study’ in 1951, where they sent a questionnaire on smoking habits to all registered British doctors. It was the first large prospective study and established a link between tobacco smoking and the risk of dying from lung cancer (Doll and Hill, 1954), as well as myocardial infarction and chronic obstructive pulmonary disease (Doll and Hill, 1956).

Evidence on the negative effects of smoking during pregnancy began to come to light towards the latter part of the 1950s and early 1960s. The evidence was pointing to smoking leading to babies being born of lower birth weight (Lowe, 1959; Simpson, 1957; Frazier et al., 1961). However, as documented by the Royal College of Physicians (RCP) the evidence at the time did not conclude that there were greater complications surrounding births.

On 7th March 1962 the RCP published “Smoking and Health”, a report highlighting the dangers of smoking for human health. The report was written with the general public in mind as opposed to the medical profession. The RCP, for the first time, put out a press release and held a press conference. The report generated attention from the media in both the press and on television.^7^ On 12th March, there was a special edition of the BBC flagship current affairs programme, *Panorama*, discussing the report, including interviews with scientists and members of the public (Berridge, 2007).^8^

The work of Doll and Hill formed the basis of the RCP’s report. Smoking was linked not only to lung cancer but also other serious conditions such as bronchitis and cardiovascular disease.^9^ The publication of the report was an important point in the history of public health: it was no longer tied to just the provision of health services. Furthermore, it led to the wider dissemination of medical research to the public, rather than it being discussed only within the medical community.

The RCP report covered the alternative hypotheses that had been put forward to attempt to explain the relationship between smoking and lung cancer, including ommitted variables that determined both smoking and lung cancer, the correlation between heavy smoking and heavy alcohol use, falling death rates of tuberculosis, reverse causality, that smoking determines the location of cancer but not the cancer itself, and motor vehicle pollution; each of these was dismissed.

The RCP report also gained traction in the US, with the US Surgeon General publishing an equivalent report in 1964. Aizer and Stroud (2010) examine the effect of the Surgeon General’s report on both maternal smoking and the health of newborns. In particular, they focus on the education-health gradient, finding that the gradient opens up after the publication of the report. This is apparent not only for smoking but they also find differences in birth weight and fetal death. James (Forthcoming) documents what happened to the education-smoking gradient in the UK after the publication of the RCP report. Using a contemporaneous survey of the general population he finds, similar to Aizer and Stroud (2010), a widening of the education-smoking gradient after the report’s publication. He also finds significant gaps in the knowledge of the dangers of smoking by education 20 years after the report was published.

The RCP report made several suggestion for ways in which the government could act to reduce smoking. There was an emphasis on education, particularly for those of school age. Other recommendations included the use of television and radio media campaigns, restricting the sale of tobacco products and smoking bans in public places, which did not come into effect in the UK until over forty years later. The report also suggested that the government provide anti-smoking clinics to help individuals quit smoking, and it called for higher (or differential) cigarette and tobacco taxes.

Restrictions on the advertising of tobacco was another recommendation of the report. Independent Television (ITV) was set up as an alternative service to the BBC in 1954, and it was funded through advertising. Although advertising was strictly controlled (Woodhouse, 1971)^10^, cigarettes were allowed to be advertised. This came to and end in 1965, when the 1964 television Act came into effect, banning all advertising of cigarettes on television. Loose tobacco and cigars could still be advertised on television, and advertising elsewhere (such as on radio or on billboards) remained permitted.

In summary, in a relatively short time since the end of the Second World War the public’s understanding of the dangers of smoking had evolved and, towards the second half of this period, the government had acted to try and reduce the harms that it caused. Alongside greater knowledge of the dangers of smoking, people now had greater access to medical care, particularly so for women and for those of lower socio-economic status. Although the information was not targeted at pregnant women, they may have been more exposed since they have regular medical appointments, and their pregnancy may make them more responsive to information.

## 3 Data

We use the UK Biobank data; a major resource that follows the health and well-being of approximately 500,000 individuals in the UK aged 40-69 between 2006-2010. They are born between 1938 and 1971, with the majority born in the late 1940s through to the early 1960s. Participants have provided information on their health and well-being, and given blood, urine and saliva samples; all participants have also been genotyped.

An advantage of the UK Biobank is that it is a very large sample of individuals for whom we observe an extensive amount of relevant health (as well as social and economic) outcomes later in life. However, it has little information on individuals’ early life environment. More specifically, the data include the location of birth, (self-reported) birth weight, an indicator for whether the participants were breastfed, and an indicator for whether the participants’ mothers smoked during their pregnancy.^11^ Our analysis explores trends in the latter, investigating how maternal smoking during pregnancy changed from the late 1930s to the early 1970s.

We are also interested in heterogeneity in the trends of maternal prenatal smoking across two dimensions. First, we explore the social gradient of maternal prenatal smoking and how this has changed over time. Since we do not observe participants’ socioeconomic status at birth, we merge in external data on the socioeconomic status at the local area-level, proxied by residents’ social class, and obtained from the 1951 Census. The 1951 Census classifies individuals into five social classes based on occupation (Register Office, 1960) and records the frequencies of each class at the Local Government District (henceforth: district) level. We exploit individuals’ location (i.e., eastings and northings) of birth to identify the district in which they were born and use that to define ‘high social class’ districts by whether the district’s proportion of residents in professional and intermediate occupations is above the median of all districts in the 1951 census year. The 1951 Census also records the number of residents by groups of educational attainment. We use an above-median proportion of residents who left education aged 20 years or older as an alternative definition of high social class districts in a robustness check.

Second, we explore heterogeneity in the trends of maternal smoking by genetic ‘predisposition’ using the molecular genetic data in the UK Biobank. See Appendix 5 for more detail on the genetic data and how our genetic variables are constructed. Whilst we do observe molecular genetic information of UK Biobank participants, we do not observe the genetic variation of their *mothers*, whose smoking decisions are our main variable of interest. However, as children inherit genetic variation from their parents, we proxy the mother’s genetics by those of the child, exploiting that children with high genetic predisposition for smoking would likely have had parents who are genetically predisposed to smoking. We use two variables capturing genetic variation in the form of single base pair substitutions called single-nucleotide polymorphisms (SNPs). First, we use a single SNP on the nicotine receptor gene CHRNA5 (RS16969968) that is well-known to correlate with how many cigarettes an individual smokes per day (see e.g., Bierut, 2010; The Tobacco and Genetics Consortium, 2010; Liu et al., 2019). For each individual, we encode the SNP by the minor allele account (taking values 0, 1, or 2), such that the number of cigarettes smoked per day increases with allele count. As an individual always receives one copy of each chromosome from their mother, two risk alleles in the child imply that the mother had at least one risk allele. Analogously, zero risk alleles imply that the mother had at most one risk allele. We exploit this to construct two subsamples to compare mothers of children with two risk alleles to those of children with zero risk alleles, thereby comparing mothers with respectively a higher and lower genetic predisposition for smoking intensity. Second, since existing genome-wide association studies have linked many SNPs to smoking, we use two polygenic indices to capture the combined effect of all such SNPs and thereby increase the predictive power for smoking behaviour compared to a single SNP. We obtain our polygenic indices from the PGI repository (Becker et al., 2021) which contains pre-computed polygenic indices for common outcomes in the UK Biobank. We focus on the polygenic indices for ‘ever smoked’ and ‘number of cigarettes per day’, allowing us to look at genetic predisposition across both the extensive and intensive margins. We standardise all polygenic indices to have mean zero, standard deviation one. Due to the child inheriting their genetic make-up from their parents, the polygenic index of the child (which we observe) will be positively correlated with the polygenic index of the mother (which we do not observe). We again exploit this to construct two subsamples of mothers with respectively higher and lower genetic ‘predisposition’ to smoking by splitting at the median of the distribution of the child’s polygenic index. Table B.1 in Appendix 5 shows the predictive power of RS16969968 and the polygenic indices for both the child’s and mother’s smoking behaviour.

We refer to genetic ‘*predisposition*’ in quotation marks. This reflects the fact that their effect is not immutable and not necessarily biological. Instead, polygenic indices can be interpreted as the best linear genetic predictor of the outcome of interest (Mills et al., 2020). In addition to potential biological effects, the association may capture gene-environment *correlation* (e.g., individuals selecting into specific environments based on their genetic variation; genetic variation invoking environmental responses). They can also capture ‘genetic nurture’: an environment shaped by *parental* genetic variation (see e.g., Kong et al., 2018). This means that polygenic indices can capture genetic as well as environmental components. Note, however, that because genes are fixed at conception, the environment cannot affect individuals’ genetic variation; there is no reverse causality. We make the following sample selection. First, to merge in data from the 1951 census, we drop individuals with missing birth co-ordinates or with co-ordinates that we cannot link to a district in England or Wales. We also drop 398 individuals born in two districts for which data on social classes is missing. This results in 404,711 individuals. Second, we drop the last two birth cohorts born in 1970 and 1971 as they are very small compared to earlier cohorts (118 and 1 individuals, respectively, compared to 1,949 born in 1969), leaving us with 404,592 individuals. We next drop those with missing data on maternal prenatal smoking, resulting in a final sample size of 348,188 individuals.

For the analyses using genetic data, we additionally drop genetic outliers, individuals of non-European genetic ancestry, and those without a polygenic index in the PGI repository. See Appendix 5 for details on this procedure. This leaves us with a final sample size of 334,573 individuals for the analyses using genetic data.

Figure 1 presents the proportion of children in our sample who indicate their mother smoked on the vertical axis by birth cohort on the horizontal axis. Maternal smoking rates increased quickly during the late 1930s and the 1940s, from approximately 18 percent in 1938 to 35 percent in 1949. In the 1950s aggregate maternal smoking rates were initially stable at approximately 35 percent and then started to fall during the second half of the decade reaching 31 percent in 1959. This downward trend continued throughout the 1960s until the end of our study period.

The vertical lines in Figure 1 mark a series of potentially important events for the evolution of (maternal) smoking, as described in the Background section. The first line indicates the end of the Second World War, the second shows 1947 which saw a large tax increase on tobacco, next is the introduction of the NHS in 1948. The fourth line is 1950 which saw the publication of Doll and Hill (1950)’s key work that began to establish the link between smoking and cancer. The next line shows the publication of the first report by the RCP on the negative health effects of smoking in 1962, and the final line shows 1965, the year when cigarette advertising was banned on television in the UK.

**Figure 1:**
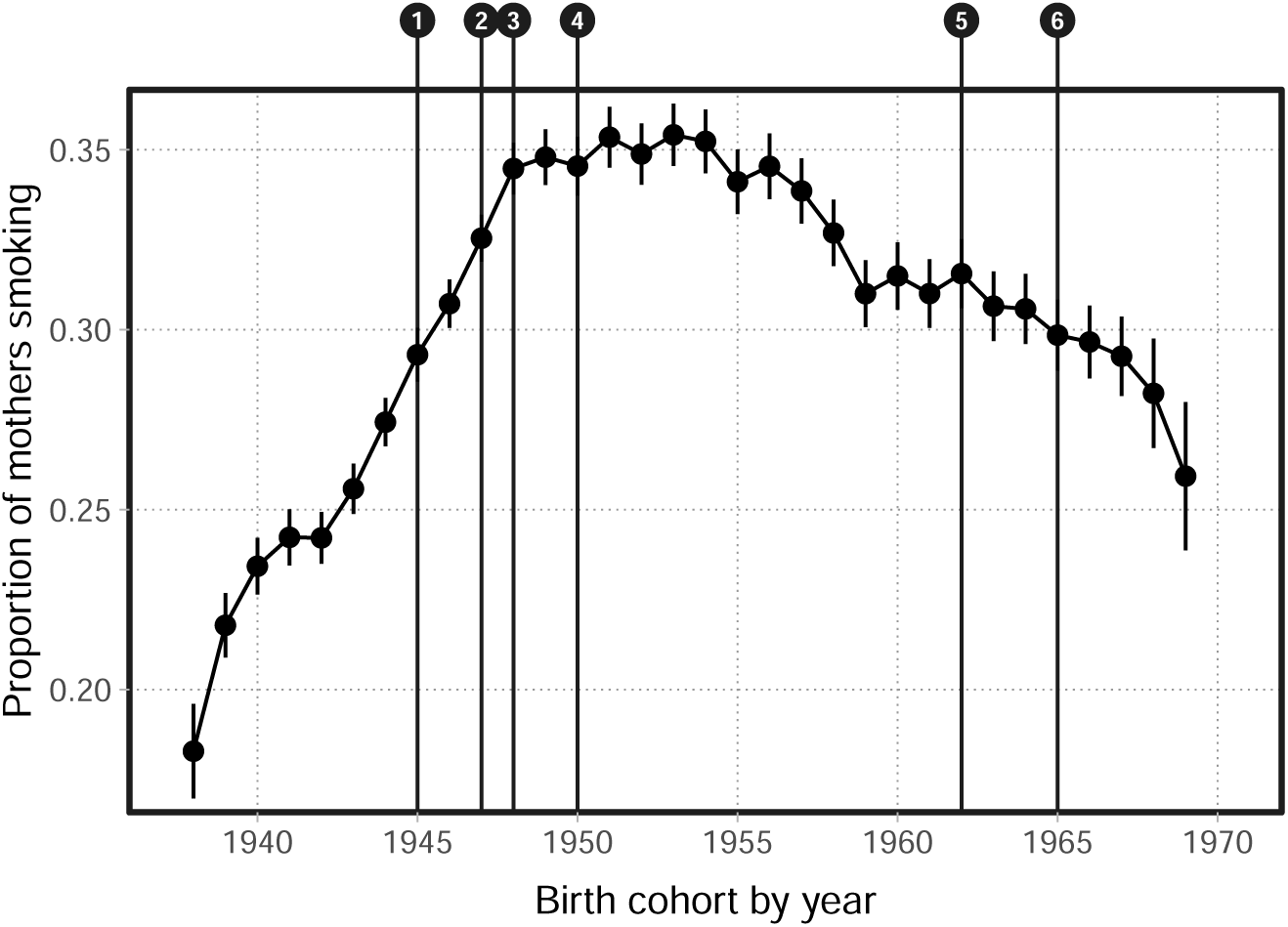
Trends in maternal prenatal smoking; 1938–1970. Figure notes: (1) WW2 ends, (2) tax hike, (3) introduction of NHS, (4) first paper on smoking and cancer, (5) RCP report, (6) ban on TV advertising of cigarettes. See the main text for details.

## 4 Heterogeneity in smoking trends

Building on the general trends shown in Figure 1 we now investigate how these trends differ by the local social class shares and genetic predisposition.

### 4.1 By district socioeconomic status

Figure 2 presents the trends in maternal prenatal smoking by the socioeconomic status of one’s district of birth, proxied by whether the proportion of district residents from high social classes in 1951 is above versus below the median proportion taken across all districts in that year. From 1938 to 1945 the levels and trends were almost identical across the two groups, starting at around 17 to 19 percent in 1938, and increasing to almost 30 percent of mothers smoking during pregnancy at the end of World War 2. After the war, a gap of around two to three percentage points opened between the two groups, with higher maternal smoking rates in low social class districts. This gap grew further after the introduction of the NHS in 1948. Following the first evidence of the harmful effects of smoking in the beginning of the 1950s, the gap increased to almost six percentage points. In 1953 the maternal smoking rate in low social class districts was 38 percent compared to 29 percent in high social class districts. Since the mid 1950s the gap between the groups has been stable, with some signs of narrowing in the last years of our study period, when the low social class group had a rate of 28 percent compared to 22 percent for the high social class group.^12^

### 4.2 By genetic variation

We now investigate the patterns in maternal smoking by children’s genetic variation as proxies for the maternal genetic ‘predisposition’ for smoking. In Figure 3 we split the sample by the individuals’ minor allele counts for SNP RS16969968 which correlates with individuals’ (and their mothers’) smoking behaviour. In contrast to Figure 2 the trends and levels are similar until the mid 1950s when the two lines start to diverge. As expected, we observe slightly higher maternal smoking rates for offspring with an allele count of two (whose mothers must have had at least one minor allele) compared to those with a count of zero. However, this gap only persists for about a decade; the levels are again indistinguishable across the two groups in the last years of our study period, potentially due to the smaller sample sizes towards the end of the observation window.^13^

**Figure 2:**
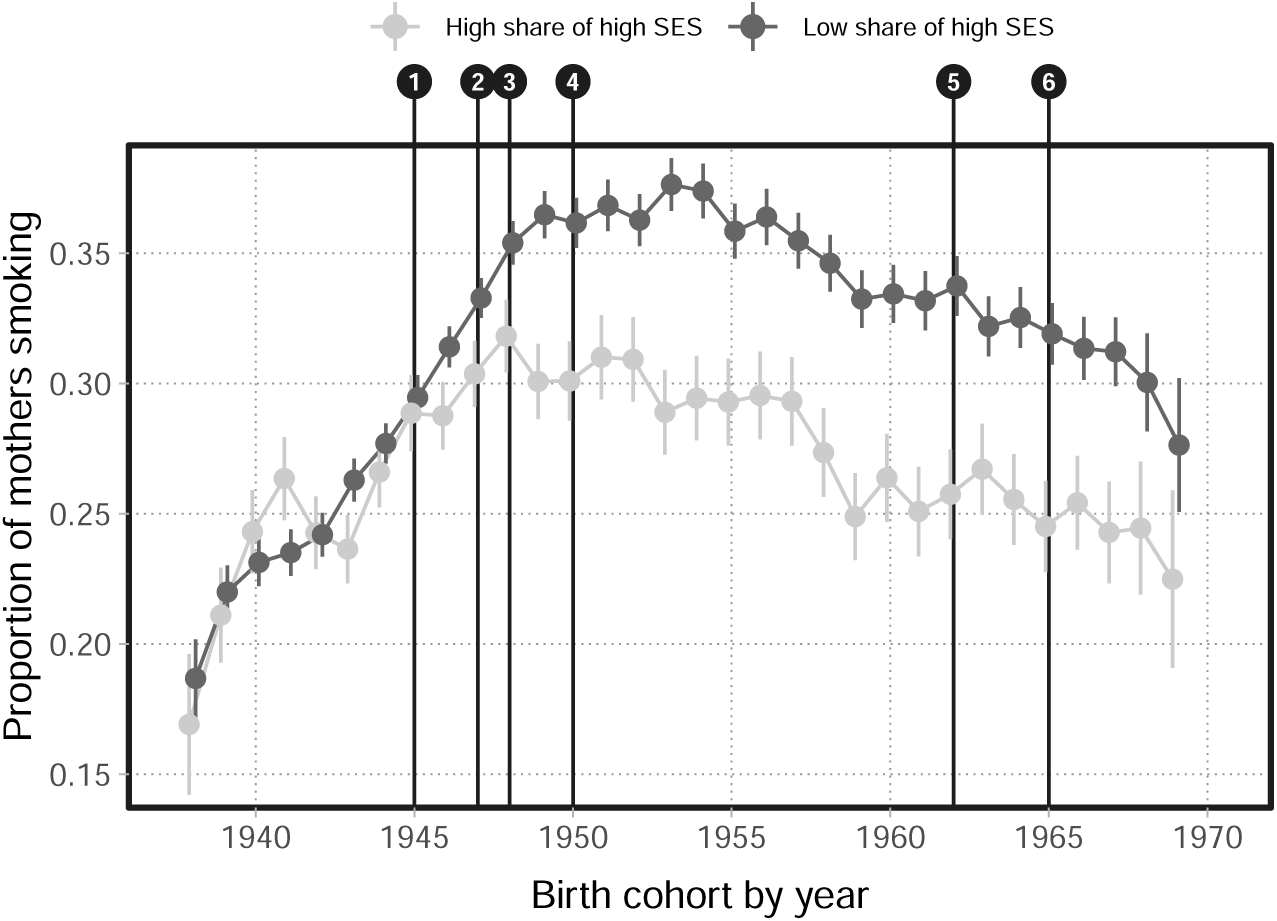
Social gradient in smoking trends; 1938–1969. Figure notes: (1) WW2 ends, (2) tax hike, (3) introduction of NHS, (4) first paper on smoking and cancer, (5) RCP report, (6) ban on TV advertising of cigarettes. Defines high SES as districts with a share of social classes 1 and 2 that are above the median taken across all districts based on the 1951 census.

**Figure 3:**
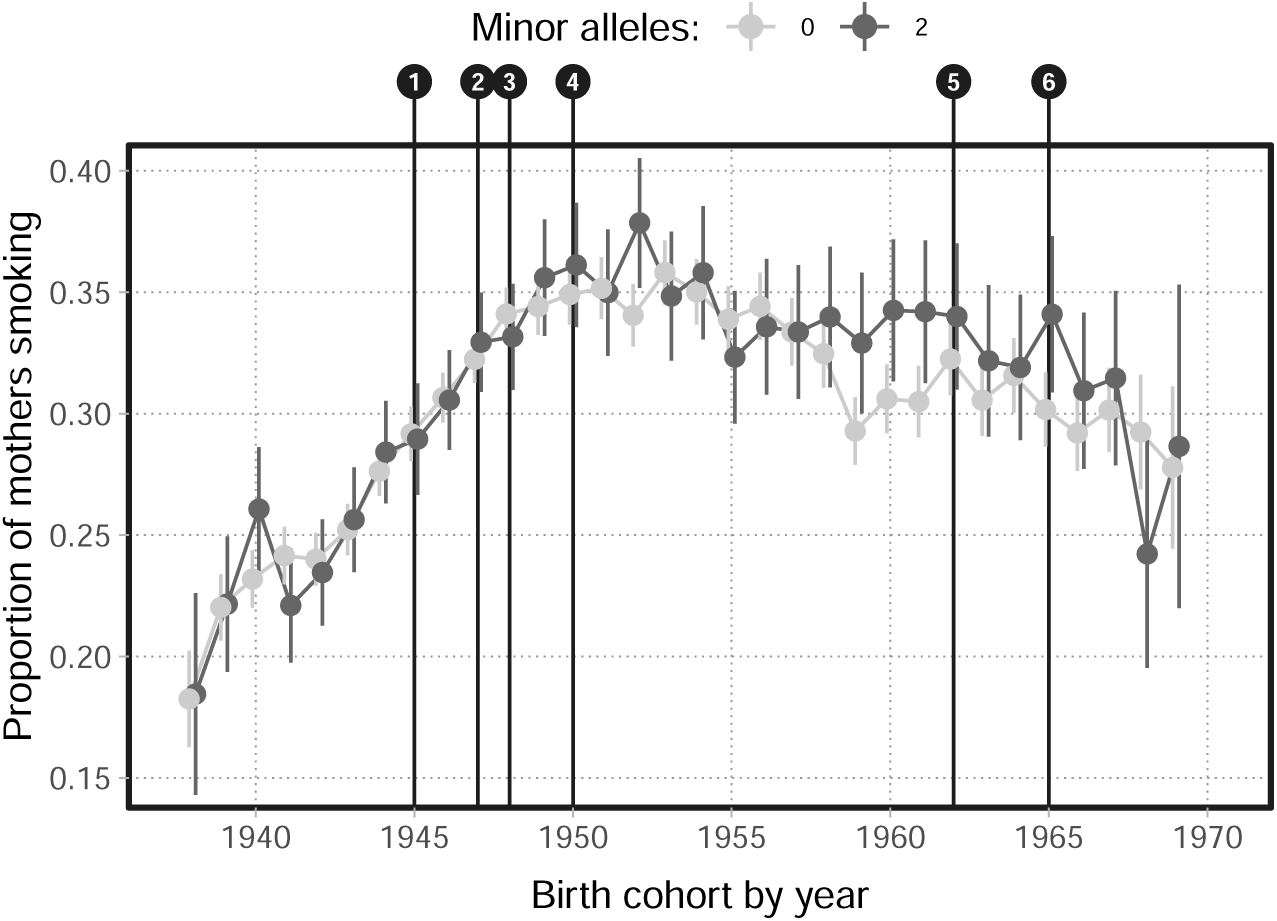
Maternal smoking by offspring’s rs16969968 minor allele count; 1938–1969. Figure notes: (1) WW2 ends, (2) tax hike, (3) introduction of NHS, (4) first paper on smoking and cancer, (5) RCP report, (6) ban on TV advertising of cigarettes.

In Figure 4 we study heterogeneity by the median split of the individuals’ polygenic index for the extensive margin of smoking (ever smoked), which correlates with the smoking statuses of their mothers (see Table B.1, Appendix 5). Compared to the earlier figures, the first difference is that the median split creates two groups with very different levels of maternal smoking throughout the study period. The above median group had a maternal smoking rate starting at about 21 percent in 1938 and increasing to more than 40 percent in the early 1950s, before it started to fall in the late 1950s. However, this fall is limited and for the last cohorts we still observe rates of about 34 percent in the above median group. The below median group started at a level of about six percentage points lower in 1938 and the rate peaked a bit earlier than for the above median group at around 30 percent in the late 1940s. This gap persists throughout the remaining study period and by the late 1960s the gap is about 11 percentage points. In Figure A.3 in the Appendix we show the patterns for the intensive margin of genetic predisposition and split the sample based on the polygenic index for ‘cigarettes per day’. We observe level differences across these two groups throughout the period, except for the first two years. Similar to our earlier findings, the gap widens and persists after World War 2.

**Figure 4:**
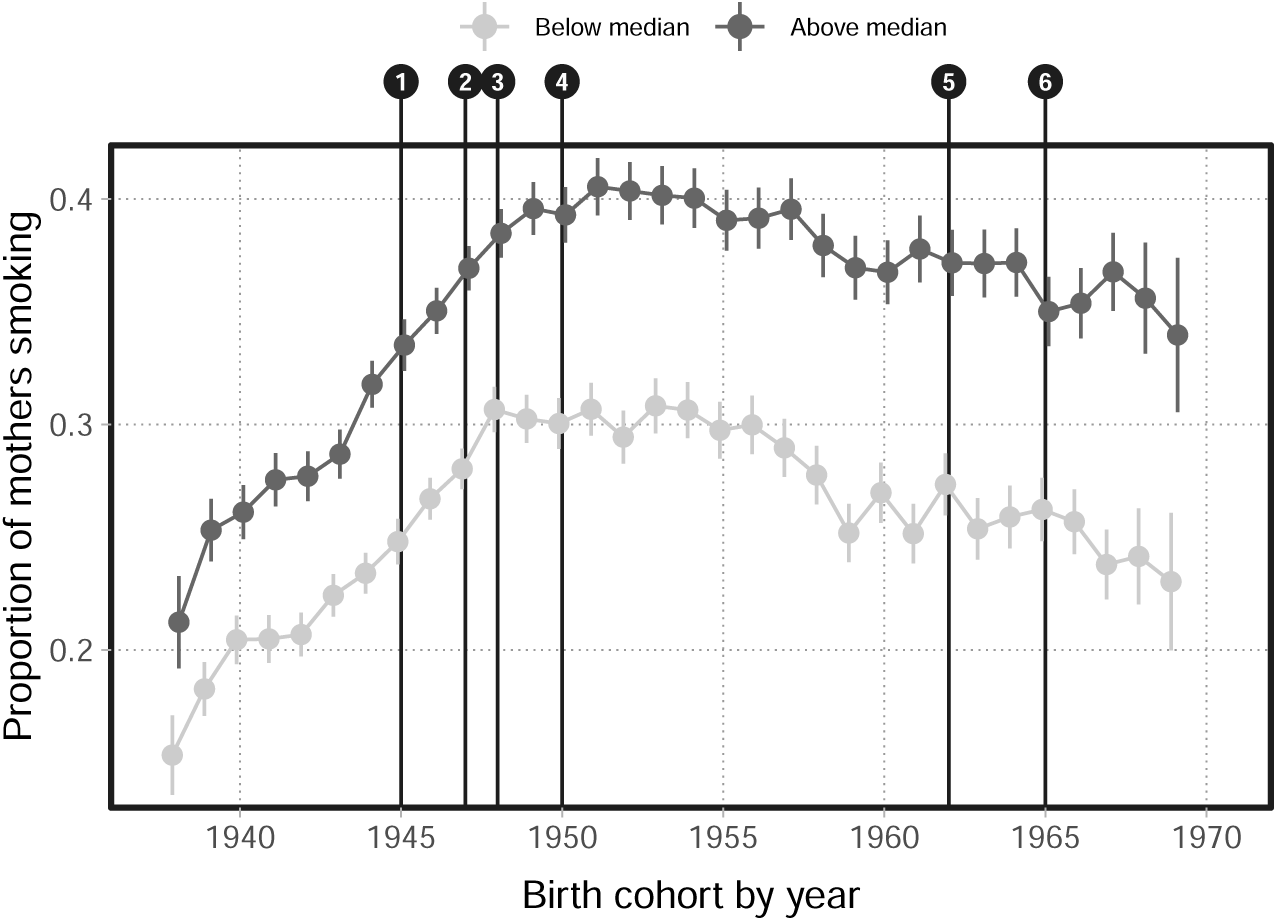
Maternal smoking by offspring’s polygenic index for ‘ever smoked’; 1938–1969. Figure notes: (1) WW2 ends, (2) tax hike, (3) introduction of NHS, (4) first paper on smoking and cancer, (5) RCP report, (6) ban on TV advertising of cigarettes.

In summary, we observe no social gradient in maternal smoking rates until after World War 2. In both low and high social class districts, we observe a smoking rate increasing from around 17-19 percent in 1938 to about 30 percent at the end of the war. After the war – coinciding with the tax changes, the introduction of the NHS, and the first evidence of the harmful effects of smoking – we observe a gap between the maternal smoking rates in high and low social class districts, with a higher rate in lower social class districts peaking at 38 percent in the early 1950s, about nine percentage points higher than for high social class districts. The gap between the groups remained throughout the period and even for children born in the last years of our study period, 1968 and 1969, we observe a gap of about five to six percentage points.

Looking at heterogeneity by polygenic index of the offspring, we observe level differences earlier on than for social class, especially when splitting the sample by genetic predisposition for ‘ever smoking’. However, also for these groups do we see a widening gap after World War 2. From 1938 up until the war, the maternal smoking rates increase in both above- and below-median polygenic index individuals, but the gap widened from about six percentage points in 1938 to around nine to ten percentage points in the early 1950s.

## 5 Conclusion

This paper presents trends in the prevalence of maternal smoking around birth in England and Wales between 1938 and 1969, and explores heterogeneities of these trends with regard to district-level social class and genetic predisposition for smoking. Maternal smoking rates increased quickly during the late 1930s and the 1940s, from approximately 18 percent in 1938 to 35 percent in 1949. Our heterogeneity analysis highlights two distinct periods during this time of substantial increase in maternal smoking. During World War 2, when tobacco was in limited supply and expensive to purchase, mothers in high- and low-social-class districts displayed very similar smoking behaviours and even the gap in smoking rates between those with an above- and below-median genetic predisposition for smoking was relatively small. This suggests that the high price of tobacco at this time may have subdued demand, preventing mothers living in disadvantaged areas (or with a high genetic predisposition) from smoking as much as they would have liked. Following the war, when these supply constraints eased, a gap in maternal smoking opens up between districts of a high and low social class and the gap by genetic predisposition widens. The large tobacco tax increase in 1947 and the NHS introduction in 1948 fall in this immediate post-war period, but due to the short time gaps between these events it is not possible to distinguish their (potential) impact from the effects of the post-war easing of tobacco supply constraints.

During the 1950s aggregate maternal smoking rates were initially stable at approximately 35 percent and then started to fall during the second half of the decade reaching 31 percent in 1959. Again, this overall trend masks some heterogeneities with regards to social class. Smoking rates among mothers from high social class districts followed a downwards trend throughout most of the 1950s; indeed the highest smoking prevalence we observe for this group was in 1948. Maternal smoking in low social class districts, on the other hand, continued to increase until 1953 and only began to fall thereafter.

This may suggest that information on the early evidence linking smoking with adverse health (in particular the link with cancer shown by Doll and Hill, 1950) may have spread differently in districts of high and low social class.

The downward trend in aggregate maternal smoking rates continued throughout the 1960s. As the RCP report was published and cigarettes were banned from being advertised on television the rates for both high- and low-class areas continued to fall as seen in our heterogeneity analysis. There are some signs of narrowing between these groups in the latter part of our analysis period. Trends for those with high and low genetic predisposition for smoking were relatively similar throughout the 1950s and 1960s, furthermore, these trends did not differ as the information on the health risks of smoking diffused through the population nor when the ban on television advertising of cigarette was introduced.

These potential links between historic policy changes, prices as well as information campaigns and the (differential) trends in maternal smoking are merely descriptive. The high frequency of relevant events during the time period we consider does not allow any causal interpretations. Existing research in health economics has, however, provided extensive causal evidence on the impact of these and other factors. Higher cigarette prices (or taxes) have been found to decrease prenatal smoking (see e.g. Evans and Ringel, 1999; Colman et al., 2003; Lien and Evans, 2005; Levy and Meara, 2006), although the reported price elasticities vary substantially between studies. While early cross-sectional research suggested smoking prevalence among youths to be strongly negatively affected by price or tax increases (e.g., Lewitt et al., 1981; Chaloupka and Grossman, 1996), more recent causal evidence shows that this price sensitivity of youth smoking is quite limited (Carpenter and Cook, 2008; DeCicca et al., 2008; Nonnemaker and Farrelly, 2011; Lillard et al., 2013; Hansen et al., 2017). Similarly, price and tax sensitivity of adult smoking behaviour has been shown to be only limited (see e.g., Farrelly et al., 2001; Forster and Jones, 2001; Sloan and Trogdon, 2004; DeCicca and McLeod, 2008; MacLean et al., 2016), especially when it comes to smoking initiation. There is furthermore evidence that adults compensate for tax increases by extracting more nicotine from each cigarette (Adda and Cornaglia, 2006) and by switching to cigarettes that are higher in nicotine and tar (Farrelly et al., 2004).

Providing information on the adverse health effects of smoking is a commonly used policy instrument. There is substantial evidence that the first large-scale reports on these adverse health consequences of smoking in the UK and US lead to a reduction in smoking and a switch to filter cigarettes with a lower tobacco content (see e.g. Sumner, 1971; Atkinson and Skegg, 1973; Warner, 1977; Schneider et al., 1981). Recent evidence shows however that this new information mainly affected smoking behaviours among more highly educated parts of the population, thus widening the educational gap in smoking (James, Forthcoming; Aizer and Stroud, 2010; de Walque, 2010). Health messages on tobacco products have been introduced in many countries since the adverse health consequence of smoking became known. Both the introduction of early text-format warnings and later pictorial warnings have been studied, with the evidence suggesting moderate reductions in smoking after mandatory warning messages were introduced (see e.g., Abernethy and Teel, 1986; Meier and Licari, 1997; Bardsley and Olekalns, 1999; Hammond, 2011; Mońarrez-Espino et al., 2014; Noar et al., 2016). Recent evidence by Kuehnle (2019) finds that pictorial warnings decreased smoking rates in Australia by encouraging smoking cessation.

Restrictions on the advertisement of tobacco products began shortly after the health consequences of smoking became widely known. Partial bans on advertisements have been shown to have limited impact on cigarette consumption in the long-run whereas comprehensive bans have been found to be more effective (see e.g. reviews by Saffer and Chaloupka, 2000; Blecher, 2008).

Smoking bans in public places have become a common policy to limit the externalities of smoking, and their causal impact on smoking behaviours have been studied e.g. by Evans et al. (1999), Carpenter (2009), Adda and Cornaglia (2010), Bitler et al. (2010), Anger et al. (2011), Carpenter et al. (2011), and Jones et al. (2015). This causal evidence is mixed and does not support a clear impact of smoking bans on smoking behaviour, but there is some evidence that the bans reduced second-hand smoke exposure. However, smoking bans have also been suggested to shift smoking to private places thus increasing second-hand smoke exposure outside of public places (Adda and Cornaglia, 2010).

Causal evidence on genetic heterogeneities relating to smoking behaviours is sparse since this area of research is still relatively young. Pereira et al. (2022) examine whether the causal effects of maternal smoking on birth weight differ by genetic ‘predisposition’ for smoking, but find no evidence of such gene-environment interactions.

Beyond the descriptive nature of our analysis, the research presented in this chapter has some further limitations. Firstly, the UK Biobank is not a representative sample of the UK population (Fry et al., 2017). Women, healthy individuals and those from less deprived areas are over-represented in the study sample.

Secondly, our data do not allow us to distinguish between trends in the general smoking behaviour of women and trends in the specific smoking behaviour during pregnancy. The maternal smoking variable used in our analysis captures both the overall prevalence of smoking as well as any changes in the likelihood to stop smoking during a pregnancy. Finally, our measure of maternal smoking is based on a 40+ year recall by the children, rather than a direct observation of smoking behaviour at the time of pregnancy. While this likely introduces some measurement error due to children not correctly recalling whether their mother smoked around the time of their birth, test-retest correlations in a subsample of participants which were asked the question a second time (at least two years after the first interview) are between 0.94 and 0.95 suggesting a strong consistency over time of the reported maternal smoking around birth. The unconventional measure furthermore allows us to study maternal smoking during a time for which no systematic data is available. It also allows us to link maternal smoking data during this period with genetic data and provides a much larger sample size than any historic survey data. As such, the data allow us to shed light on the stark trends in maternal smoking around pregnancy during the war and post-war reconstruction period, and highlight the substantial heterogeneities in smoking behaviour during this time.

## Data Availability

This work is based on data from the UK Biobank which are accessible to researchers online by application and payment of a fee. This work is also based on data provided through www.VisionofBritain.org.uk and uses historical material which is copyright of the Great Britain Historical GIS Project and the University of Portsmouth.

https://www.ukbiobank.ac.uk

http://www.VisionofBritain.org.uk

### Appendix A: Additional tables and figures

**Figure A.1:**
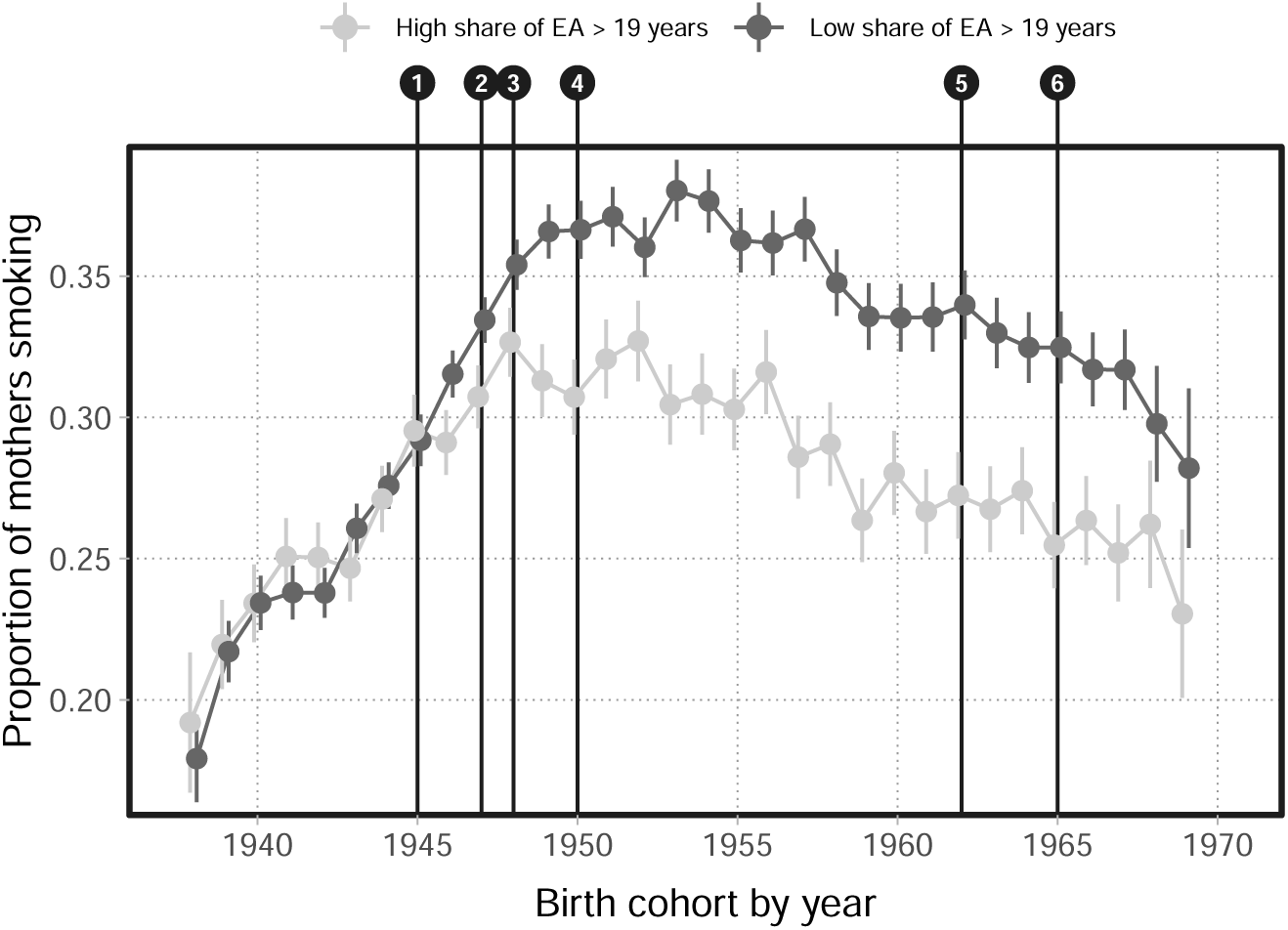
SES defined according to frequency of educational attainment in 1951. Figure notes: (1) WW2 ends, (2) tax hike, (3) introduction of NHS, (4) first paper on smoking and cancer, (5) RCP report, (6) ban on TV advertising of cigarettes. Defines high education areas as districts with an above-median share of those who left education aged 20 years or older, taken across all districts based on the 1951 Census.

**Figure A.2:**
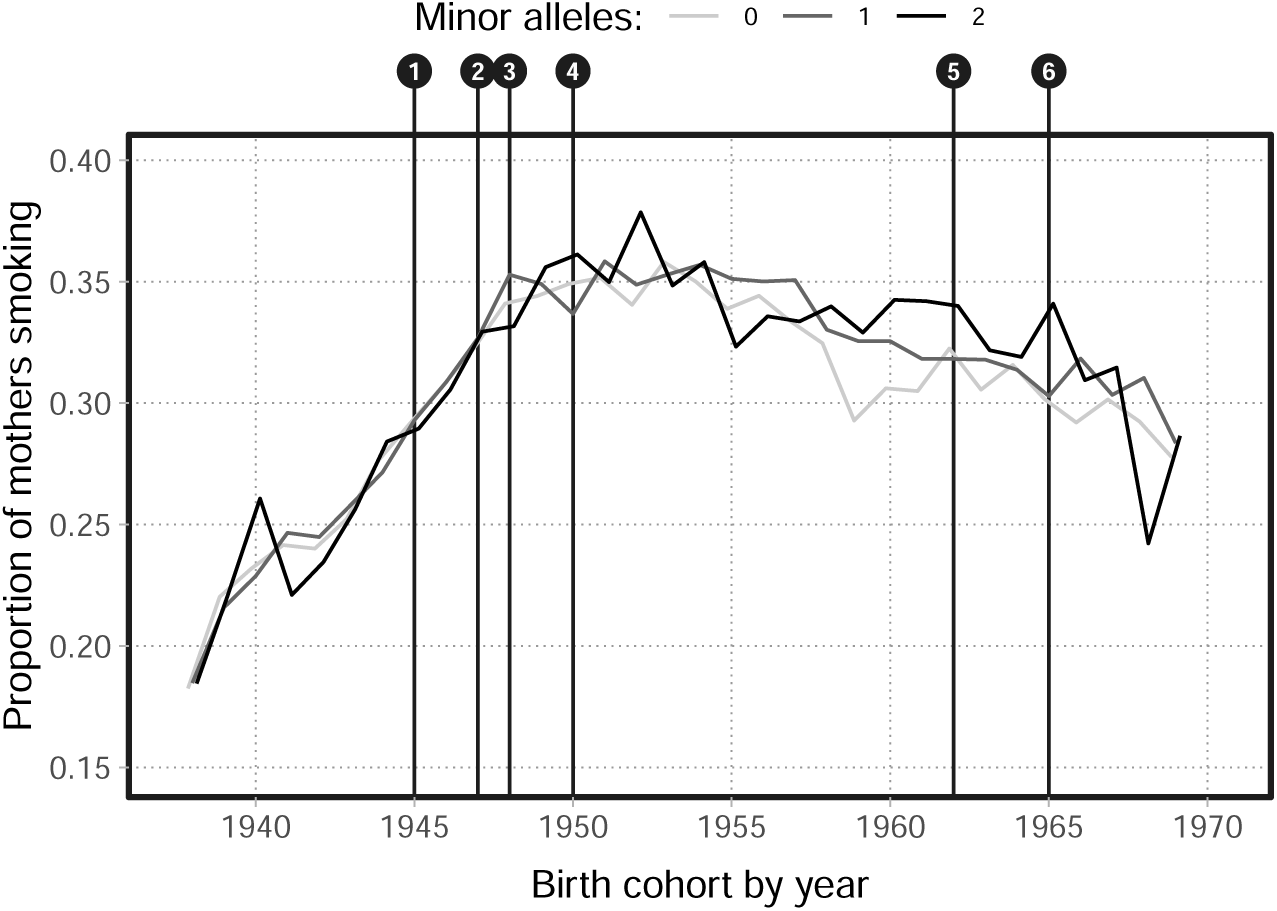
Maternal smoking by offspring’s RS16969968 minor allele count; 1938–1969. Figure notes: (1) WW2 ends, (2) tax hike, (3) introduction of NHS, (4) first paper on smoking and cancer, (5) RCP report, (6) ban on TV advertising of cigarettes.

**Figure A.3:**
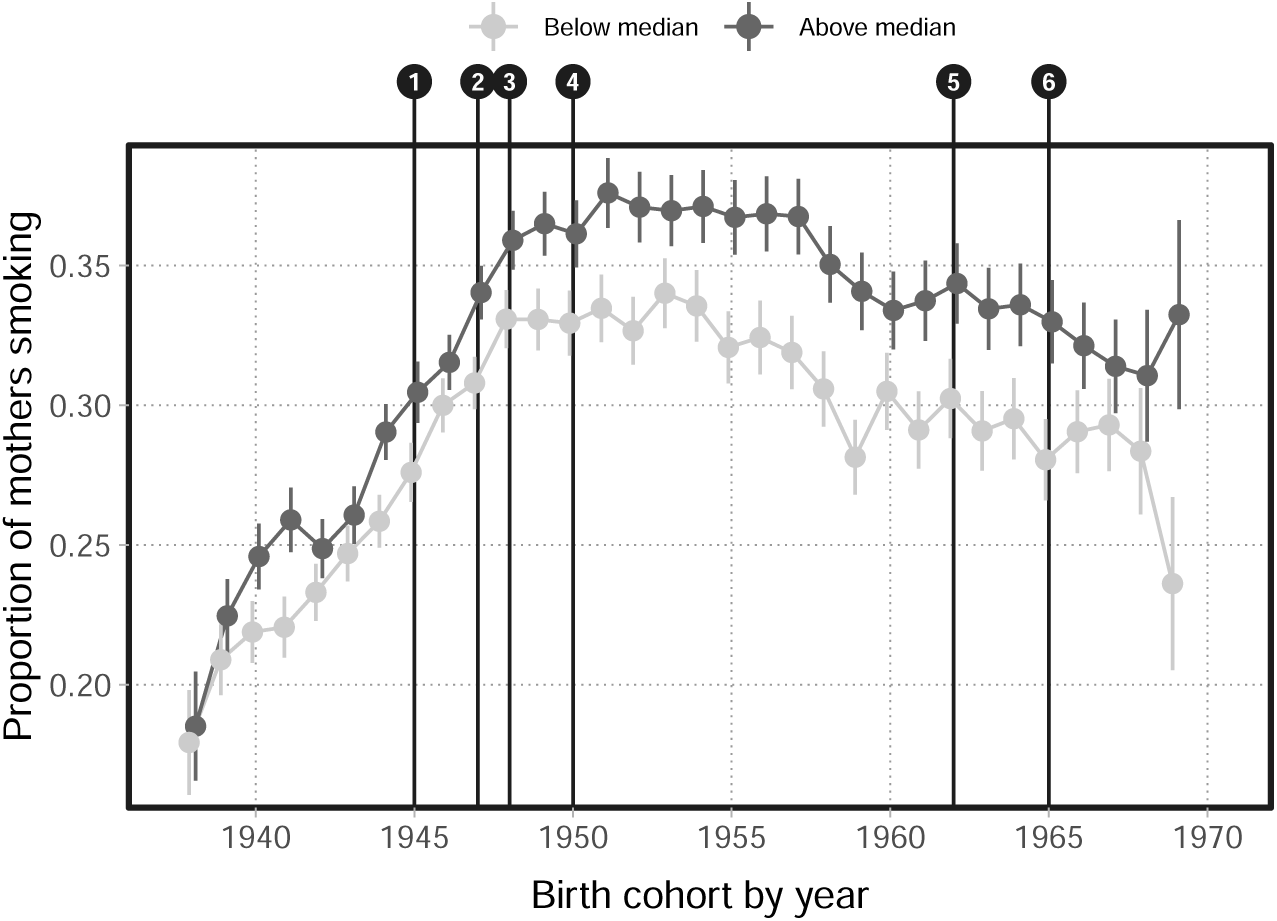
Maternal smoking by offspring’s polygenic index for ‘cigarettes per day’; 1938–1969. Figure notes: (1) WW2 ends, (2) tax hike, (3) introduction of NHS, (4) first paper on smoking and cancer, (5) RCP report, (6) ban on TV advertising of cigarettes.

### Appendix B: Genetics

Humans have 23 chromosome pairs in every cell apart from sex cells. Each pair contains a maternal and paternal copy inherited from respectively the mother and the father. A single chromosome consists of a double-strand of deoxyribonucleic acid (DNA) containing a large number of ‘base pairs’: pairs of nucleotide molecules (referred to as the ‘letters’ A (adenine) that binds with T (thymine), and G (guanine) that binds with C (cytosine)) that together make up the human genome. Across a population there will be locations in the genome where a single base pair has been replaced by a different one. Such variation is known as a Single Nucleotide Polymorphism (SNP, pronounced ‘snip’) and is the most commonly studied type of genetic variation. When there are two possible base pairs at a given location (i.e., two alleles), the most frequent base pair is called the major allele, while the less frequent is called the minor allele. As humans have two copies of each chromosome, any given individual can have either zero, one, or two copies of the minor allele at a given location.

To identify specific SNPs that are robustly associated with a particular outcome of interest, so-called Genome-Wide Association Studies (GWAS) relate each SNP to the outcome. As there are more SNPs than individuals, the SNP effects cannot be identified in a multivariate regression model. Instead, a GWAS runs a large number of univariate regressions of the outcome on each SNP. These analyses have shown that most outcomes of interest in the social sciences are ‘polygenic’: they are affected by a large number of SNPs, each with a very small effect. To increase the predictive power of the SNPs, it is therefore custom to aggregate the individual SNPs into so-called polygenic indices, by constructing a weighted sum of the minor allele counts at each SNP, where the weight is the effect size obtained from a GWAS (after additionally accounting for correlations between SNPs).

We construct two measures of genetic predisposition to smoking. First, we focus on the single SNP RS16969968 on chromosome 15 (in the UK Biobank we identify this SNP by position 78590583 [GRCh37/hg19]), and construct a variable counting the number of risk alleles each individual has at this location. We code A/T (the minor allele) as the risk allele such that smoking propensity increases with risk allele count. Second, we also use two polygenic indices from the PGI repository (Becker et al., 2021) to capture genetic predisposition for smoking aggregated across many SNPs. The PGI repository contains pre-computed polygenic indices for common outcomes in the UK Biobank. Becker et al. (2021) maximize the predictive power of the indices by meta analysing several large datasets. We focus on the indices for ‘ever smoked’ and ‘number of cigarettes per day’, allowing us to look at genetic predisposition across both the extensive and intensive margins. Becker et al. (2021) details the construction of the polygenic indices.^14^

To verify the predictive power of RS16969968 and the polygenic indices, we use a linear regression model to test how well they predict smoking outcomes in our UK Biobank sample. We QC the UK Biobank genetic data and identify individuals of European genetic ancestry using the procedure described in Elsworth et al. (2019). Our regressions control for sex and the first 20 genetic principal components, and we standardize the polygenic indices to have zero mean and unit variance in the sample. Since Becker et al. (2021) constructed the polygenic indices using a three-fold sample split with overlap in the GWAS discovery sample, we note that the standard errors in the regressions for the polygenic indices will be underestimated. However, the impact of this is negligible in our main analyses as we only use the polygenic indices to create an above/below median split of genetic predisposition.

Table B.1 reports the regression results for the three genetic variables (panels) and four different smoking outcomes (columns) from the UK Biobank. We consider the smoking status of the mother around the time of birth (Column 1), the child ever having smoked (Column 2), and the child’s number of cigarettes smoked per day for current (Column 3) and previous smokers (Column 4). Note that the number of cigarettes smoked per day is only available for current and past smokers, so the sample sizes are smaller for these outcomes. For each regression Table B.1 also shows the incremental *R*^2^, defined as the increase in *R*^2^ when the genetic variable is included as a covariate.

Starting with maternal smoking in Column 1, we find that all three of the child’s genetic variables predict whether the mother was smoking around the time of birth, giving supporting evidence to our use of the child’s genetics as a proxy for the mother’s. In Column 2-4, we examine how the child’s genetic variables predict the *child’s* outcomes, and we find that both polygenic indices positively predict ‘ever smoking’ as well as the number of cigarattes per day for both current and past smokers. Similarly, we see that RS16969968 also positively predicts number of cigarettes per day, but shows a negative association with ‘ever smoked’. We interpret this as RS16969968 being predictive of smoking intensity, rather than being predictive of the extensive margin, capturing whether or not the individual smokes.

**Table B.1:**
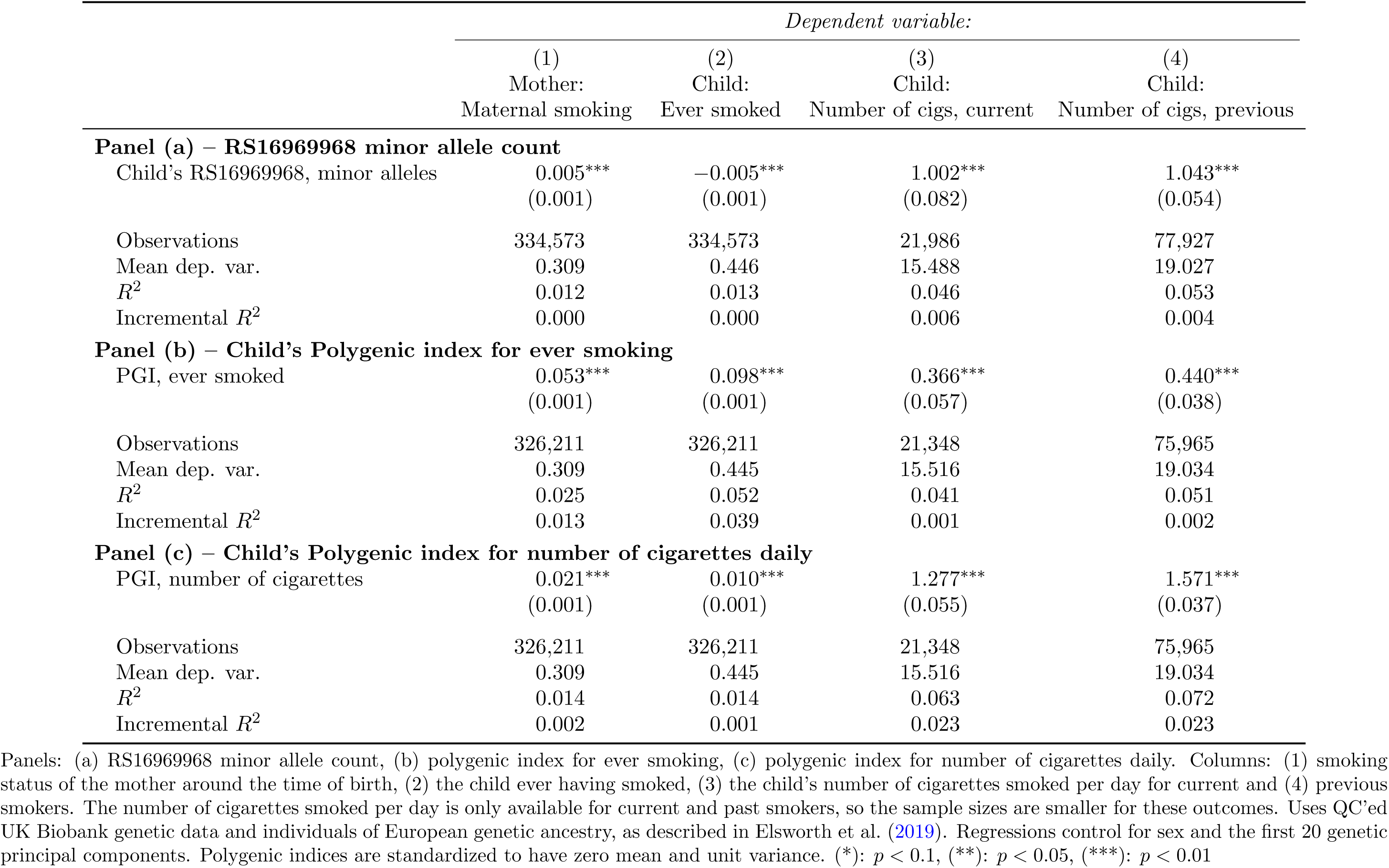
Predictive power of the genetic variables.

## Footnotes

1 Given the difficulty of accounting for the endogeneity of maternal smoking, the empirical specifications used range from simple linear regressions to instrumental variable regressions to mother fixed effects and matching approaches.

2 Indeed, adult outcomes are shaped by the early life environment, including nutrition, alcohol, smoking, disease, mental health, economic conditions, pollution, and so on (for reviews, see e.g., Almond and Currie, 2011a,b; Almond et al., 2018).

3 Hansard, 15 April 1947

4 Hugh Dalton’s Budget, 1947

5 To ease the impact on old-age pensioners, MPs urged Dalton to provide OAPs access to cheap tobacco. The government agreed and from October 1947, OAPs who could prove they were ‘habitual smokers’ could purchase a limited amount of tobacco each week at a reduced price (Singleton, 2023).

6 “Your New National Health Service”, 5 July 1948.

7 See the archive footage from the British Broadcasting Corporation (BBC) on the report.

8 Although the relationship between parental smoking and children’s birthweight was discussed, it was only afforded one paragraph in the 70 page pamphlet.

9 Within the RCP there was a discussion that suggested the report should also espouse the dangers and risks of air pollution. This did not happen in order to prevent the message that smoking led to lung cancer and other health problems being watered down (Berridge, 2007).

10 Six minutes per hour were allowed, sponsorship of programmes was not permitted, and the scripts and advertisements themselves were reviewed by representatives of the “Independent Television Authority” (ITA).

11 The exact question in the UK Biobank is “Did your mother smoke regularly around the time when you were born?”

12 In Figure A.1 in the Appendix we split the sample by district shares of educational attainment. The pattern is similar to Figure 2 with low education districts mimicking the low social class districts, and high education districts the high social class districts.

13 In Figure A.2 in the Appendix we additionally include offspring with an allele count of 1 in the analysis.

14 For background on how to interpret polygenic indices and on the PGI repository more generally, see also the FAQs for Becker et al. (2021) at https://www.thessgac.org/faqs.

## Notes

### Competing Interest Statement

The authors have declared no competing interest.

### Funding Statement

We gratefully acknowledge financial support from NORFACE DIAL (462-16-100) and the European Research Council (ERC) under the European Union's Horizon 2020 research and innovation programme (grant agreement No. 851725).

### Author Declarations

UK Biobank has received ethical approval from the National Health Service Northwest Centre for Research Ethics Committee (11/NW/0382) and has obtained informed consent from its participants.

